# Using Latent Class Analysis to Identify Subgroups of Post-Operative Older Adults

**DOI:** 10.1101/2025.01.03.25319954

**Authors:** Kevin McLaughlin, Amie Bettencourt, Daniel Young, Erik Hoyer, Michael Friedman, Elizabeth Colantuoni, Lee A. Goeddel, Pedro Gozalo

**Affiliations:** Johns Hopkins University School of Medicine, Department of Physical Medicine and Rehabilitation; Johns Hopkins University School of Medicine, Department of Psychiatry and Behavioral Sciences; University of Nevada Las Vegas, Department of Physical Therapy; Johns Hopkins Bloomberg School of Public Health, Department of Biostatistics; Johns Hopkins University School of Medicine, Department of Anesthesiology and Critical Care Medicine; Brown University School of Public Health, Department of Health Services, Policy and Practice

## Abstract

**Objective:** Identify subgroups of postoperative older adults using electronic health record data.

**Summary of Background Data:** Postoperative older adults represent a vulnerable population who may benefit from tailored postoperative care pathways. Identifying clinical subgroups can inform the development of these pathways.

**Methods:** Retrospective cohort study of postoperative adults ≥65 years (N=2,036) from a single healthcare system. Latent class analysis was used to identify patient subgroups based on measures of frailty, mobility, activities of daily living, and general health status. Hospital outcomes were described among each subgroup, including extended lengths of stay (LOS) (>0.5 SD beyond mean LOS by surgical category), discharge disposition (i.e., home versus non-home discharge), and utilization (weekly visit frequency) of physical therapy (PT) and occupational therapy (OT).

**Results:** We identified 3 subgroups that we labeled Low Frailty-High Mobility (LF-HM), High Frailty-Low Mobility (HF-LM), and Low Frailty-Low Mobility (LF-LM), representing 15.3%, 27.6%, and 57.1% of the cohort, respectively. Discharge to home was highest among the LF-HM group (99%), followed by LF-LM (96%), and HF-LM (77%). Extended LOS was most common among the HF-LM group (27%), followed by LF-LM (18%), and LF-HM (6%). PT and OT visit frequencies were highest in the HF-LM group followed by the LF-LM and LF-HM groups.

**Conclusions:** This study identified 3 subgroups of postoperative older adults using routinely collected patient data. These groups may help to identify patients with increased odds of non-home discharge, extended LOS, and higher utilization of PT and OT and may inform the development of tailored postoperative care pathways for older adults.

## Introduction

More than half of the 20 million surgeries performed each year in the US are for adults who will require an inpatient hospital stay.^1^ Among these, older adults are at increased risk for extended hospital lengths of stay (LOS) and discharge to post-acute care facilities,^2–7^ which conflict with patient preferences and contribute to increased cost of care.^8–10^ Enhanced recovery after surgery (ERAS) pathways have been shown to improve clinical outcomes among post-operative older adults.^11–15^ However, ERAS pathways typically include additional interventions or services (e.g., medication rehabilitation services), with associated costs, and not all patients are equally likely to benefit from these services. There is a need to better identify older adult patients at risk of poor post-operative outcomes to better target resources and tailor postoperative care pathways to the needs of individual patients.

Previous studies have identified individual patient-level factors associated with post-operative outcomes. For example, studies have found that pre-operative frailty is associated with increased risk for loss of independence following surgery, as well as longer LOS, among older adults.^7,16^ Another study found that patients with lower levels of mobility one day after gastrointestinal surgery were more likely to experience an extended LOS.^17^ Additional studies have observed associations between age, gender, body mass index, pre-operative function and American Society of Anesthesiologists (ASA) rating with post-operative outcomes following surgery.^2,6,18,19^ However, few studies have examined preoperative and postoperative patient data in combination, which is likely to account for variability in patient outcomes better than preoperative or postoperative data alone.

Latent class analysis is a statistical approach that is increasingly being used to identify subgroups of patients within heterogenous clinical populations who have similar patterns of clinical outcome techniques.^20–23^ Identifying subgroups in this manner is commonly used to inform the development of tailored or “precision” treatment approaches.^20,23–26^ The purpose of this study was to identify subgroups of post-operative older adults using a latent class analysis of routinely collected patient-level data from a large academic health care system. We also examined differences in key post-operative outcomes among those in each of the identified latent subgroups.

## Methods

We utilized a retrospective cohort design to identify latent subgroups of older adults admitted to the hospital following surgery and to examine differences in hospital outcomes among those in each latent subgroup. Data for this study were collected during routine care encounters and extracted from the electronic health record (EHR) for analysis.

### Patient Cohort

We included older adults (≥65) hospitalized at Johns Hopkins Hospital or Johns Hopkins Bayview Medical Center following inpatient surgery between September 2016 and March 2020. Eligible patients included those having any of the following surgical procedures: abdominal surgery, gynecologic surgery, pancreatic surgery, head & neck surgery, neurosurgery, orthopaedic surgery, plastic surgery, spine surgery, thoracic surgery, urologic surgery, or vascular surgery. Individuals undergoing outpatient surgery were not included in our cohort.

### Latent Class Indicators

Based on previous evidence and standard data collection practices at our institution, we utilized patient age, general health status, frailty, mobility, and activities of daily living to identify latent subgroups of older adults after surgery.^2,6,7,16–19^ General health status was measured prior to surgery by the anesthesiology team using the American Society of Anesthesiologists Physical Status Classification System (ASA PS Classification), a method used to succinctly assess and communicate patients’ pre-anesthesia medical comorbidities.^27^ ASA PS Classification scores range from 1 (normal health patient) to 6 (brain-dead). Frailty was measured at pre-operative appointments by nursing using the Edmonton Frail Scale (EFS), a valid and reliable measure of frailty with scores ranging from 0-17 and higher scores indicative of greater frailty.^28,29^ Patient mobility function was assessed using the Activity Measure for Post-Acute Care (AM-PAC) Inpatient Basic Mobility “6-Clicks” Short Form.^30,31^ Patient function in activities of daily living was measured using the AM-PAC Inpatient Daily Activity “6-Clicks” Short Form. Both AM- PAC short forms include 6 items scored by clinicians on a scale of 1-4 with lower scores indicating greater impairment. Raw scores were converted to t-scores and used in all analyses; these t-scores range from 16.6 to 57.7.^32^ In our hospitals, AM-PAC short forms are scored daily by nursing and at all PT visits (mobility short form) and OT visits (daily activity short form). Previous studies have shown high interrater reliability across and within disciplines for these measures.^33,34^ We utilized the first observed AM-PAC mobility and daily activity scores recorded by either discipline following surgery for analysis.

### Hospital Outcomes

Hospital outcomes for this study included LOS, discharge disposition, and utilization of physical therapy (PT) and occupational therapy (OT) services. LOS was categorized as an extended LOS, defined as a LOS (days) that was ≥0.5 standard deviations above the mean LOS for each surgical category, or a non-extended LOS, defined as any LOS less than the extended LOS definition. Discharge disposition was categorized as home discharge or non-home discharge (e.g., skilled nursing facility, inpatient rehabilitation facility). PT and OT utilization were examined separately based on weekly visit frequency, which was identified based on the number of PT or OT visits received, divided by patients’ LOS, and multiplied by 7.

### Analysis

Descriptive statistics were used to describe the sample. LCA was used to identify classes based on the continuous variables: age, AM-PAC mobility and AM-PAC daily activity scores, and dichotomized variables: ASA ratings, EFS total score, and individual EFS items (i.e., general health status, functional dependence, functional performance). ASA ratings were dichotomized to those patients who were healthy or with mild systemic disease (i.e., scores of 1-2) or patients with severe disease (i.e., scores of 3 and above). The general health status item on the EFS was dichotomized to those patients in good to excellent health or patients in fair to poor health. The functional dependence criterion of the EFS was dichotomized to those who required help in 0-1 activities or those who required help in 2 or more activities. The functional performance criterion of the EFS was dichotomized to those who completed the timed up and go test in 0-10 seconds or for those who completed it in 11 or more seconds. All LCA models were estimated using Mplus, Version 8.6.^35^ Fit statistics, classification accuracy and relative class size were considered in selecting the best fitting latent class model.

Model fit was measured using the Bayesian information criterion (BIC) and the sample size-adjusted BIC (aBIC).^36^ For the BIC and aBIC, smaller values indicate better model fit. The Vuong-Lo-Mendell-Rubin Likelihood Ratio Test (VLMR-LRT) was used to compare the relative fit of a model with *k* classes to a model with one fewer class.

Significant p values for the VLMR-LRT indicate that the latent class solution with fewer classes should be rejected in support of the solution with more classes.^36^ Entropy, ranging from 0 to 1, was also examined with larger values indicating better classification accuracy. Finally, class sizes were examined for each solution as simulation studies indicate that small or uncommon classes can be difficult to identify reliably and it is important to avoid over extracting classes.^36^

Once the unconditional latent class model was established, we used multinomial logistic regression to examine class differences in two dichotomous (i.e., discharge disposition, extended length of stay) and two continuous hospital outcomes (i.e., PT and OT weekly frequencies).

Next, the auxiliary function with Bolck, Croon and Hagenaars (BCH) weights was used to examine differences in the prevalence of these outcomes across classes. In the BCH approach, weights were applied to individual participants based on their posterior probabilities of class membership and an overall test of equality as well as pairwise tests of prevalence differences across classes using one degree of freedom were conducted.^35,37^

### Ethical Statement

This study was acknowledged as exempt by the Johns Hopkins Medicine Institutional Review Board (IRB00337243). Data were collected for all participants under a waiver of consent.

## Results

We identified 2,036 older adults (≥65) hospitalized after eligible surgical procedures between September 2016 and March 2022. Our cohort was 45% female and the majority of patients were White (76.6%) or Black (16.1%). The average length of stay among included patients was 4.4 days (SD: 4.4). Length of stay varied by surgical category, from 2.8 days (SD: 3.3) in urologic surgery to 8.5 days (SD: 5.7) in pancreatic/hepatobiliary surgery. The most common surgical categories were abdominal surgery (21.2%) and urologic surgery (20.3%) (Table 1).

**Table 1.** Cohort Description.

|  |  |
| --- | --- |
| **Age**, mean (SD) | 73.0 (5.9) |
| **Female**, No. (%) | 917 (45.0) |
| **Race**, No. (%) |  |
| White | 1559 (76.6) |
| Black | 328 (16.1) |
| Asian | 70 (3.4) |
| Other | 79 (3.9) |
| **Ethnicity**, No. (%) |  |
| Hispanic | 46 (2.3) |
| Non-Hispanic | 1973 (96.9) |
| Other/unknown | 17 (0.8) |
| **Length of stay (days)**, mean (SD) | 4.4 (4.4) |
| **Surgical specialty**, No. (%) |  |
| Abdominal | 432 (21.2) |
| Gynecology | 89 (4.4) |
| Hepatobiliary/pancreatic | 239 (11.7) |
| Head and neck | 59 (2.9) |
| Neurosurgery | 194 (9.5) |
| Orthopaedics | 100 (4.9) |
| Plastics | 65 (3.2) |
| Spine | 287 (14.1) |
| Thoracic | 99 (4.9) |
| Urology | 414 (20.3) |
| Vascular | 58 (2.8) |
| **Total**, No. | 2036 |

For our LCA, comparison of the fit statistics, classification quality, and class size for solutions specifying one to eight classes suggested several reasonable models (see Table 2). Although the eight-class model had the lowest BIC and aBIC values, the scree plot indicated that minimal benefit of extending the model beyond five-or six-classes (visualized as the line flattening between the models with five or six classes indicating minimal decrease in BIC and aBIC for models with more than 6 classes). The VLMR indicated that a four-class model fit the data better than a three-class model and a five-class model fit better than a four-class model but adding a sixth class did not improve model fit. However, the smallest class in the four-class model included only 2-3% of the sample (33-57 people) suggesting that these patients were outliers on each of the measures, and not a particularly clinically meaningful class. Based on all of these considerations, we selected the three-class model as the best fitting model and further analyses focused on that solution.

**Table 2.**
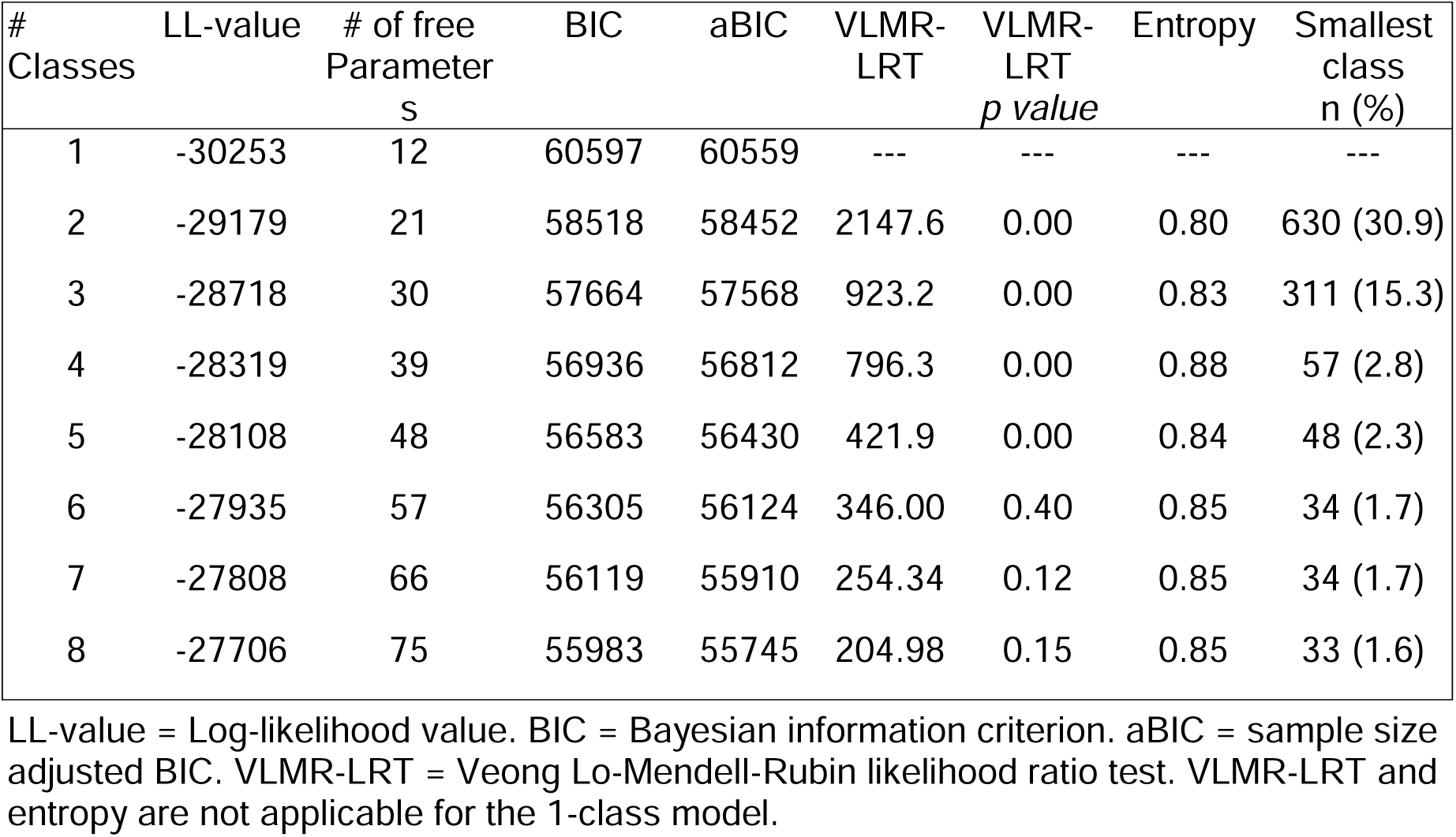
Fit Indices for Latent Class Models.

Based on the characteristics of the members of each of the identified classes, we labeled the 3 identified classes as the Low Frailty-High Mobility (LF-HM) class (15.3%), the High Frailty-Low Mobility (HF-LM) class (27.6%), and the Low Frailty-Low Mobility (LF-LM) class (57.1%) (Table 3). Age was similar between the LF-HM and LF-LM classes (71.6 and 72.5 years, respectively), but was higher in the HF-LM class (75.0 years). Frailty scores were similar in the LF-HM class (2.6) and LF-LM class (2.2) but were notably higher among those in the HF-LM class. Mean AM-PAC mobility scores were highest in the LF-HM class (54.2), followed by the LF-LM class (41.6), and HF-LM class (37.7). AM-PAC daily activity scores followed a similar pattern with mean scores of 54.8, 40.7, and 37.5 in the LF-HM, LF-LM, and HF-LM classes, respectively. ASA also differed by class with the HF-LM class having the highest probability (0.87) of receiving an ASA rating of 3 or higher and the LF-HM and LF-LM having similar but lower probabilities of receiving an ASA rating of 3 or higher (0.64 and 0.67, respectively). Individual EFS items also differed by class. Specifically, individuals in the HF-LM had a much higher probability of having fair to poor health status, being functionally dependent, and requiring more than 10 seconds to complete the up and go test as compared to the LF-HM and LF-LM classes.

**Table 3:** Description of Identified Classes and Outcomes of Class Membership (N=2036)

| Class Indicators |  | Low Frailty-High Mobility<br>(15.3%) | High Frailty-Low Mobility<br>(27.6%) | Low Frailty-Low Mobility<br>(57.1%) |
| --- | --- | --- | --- | --- |
|  | Age | 71.59 | 75.03 | 72.46 |
|  | EFS Total Score, mean | 2.57 | 6.46 | 2.20 |
|  | AMPAC Mobility t-score, mean | 54.24 | 37.68 | 41.59 |
|  | AMPAC Daily activity t-score, mean | 54.78 | 37.53 | 40.68 |
|  | ASA Rating 1-2, (%) | 0.36 | 0.13 | 0.33 |
|  | ASA Rating 3-4, (%) | 0.64 | 0.87 | 0.67 |
|  | Health status -0, (%) | 0.75 | 0.27 | 0.79 |
|  | Health status -1-2, (%) | 0.25 | 0.73 | 0.21 |
|  | Functional dependence -0, (%) | 0.96 | 0.49 | 0.98 |
|  | Functional dependence -1-2, (%) | 0.05 | 0.51 | 0.02 |
|  | Functional performance -0, (%) | 0.70 | 0.15 | 0.74 |
|  | Functional performance -1-2, (%) | 0.30 | 0.85 | 0.26 |
| Hospital Outcomes | Discharge to home, % (SE) | 99 (1)† | 77 (2)† | 96 (1)† |
|  | Extended length of stay*, % (SE) | 6 (2)† | 27 (2)† | 18 (1)† |
|  | Physical therapy visits (per week), mean (SE) | 0.3 (0.07)† | 2.1 (0.10)† | 1.2 (0.06)† |
|  | Occupational therapy visits (per week), mean (SE) | 0.1 (0.04)† | 1.1 (0.07)† | 0.5 (0.04)† |

N=2,036.
*Note:* For categorical hospital outcomes, values by class represent proportions. For continuous outcomes values represent means.
†Significantly different compared to other classes in row ( $p < 0.001$ )
\*Represents a length of stay $\geq 1$ SD above mean length of stay (days), by surgical category

Class membership varied by surgical category (Figure 1). For example, the LF-HM class was most represented among those undergoing head & neck surgery (32%) and was least represented among those undergoing hepatobiliary/pancreatic surgery (2%). The HF-LM class was most represented among those undergoing vascular surgery (50%) and least represented by those undergoing urologic surgery (12%). Finally, the LF-LM class was most represented by those undergoing hepatobiliary/pancreatic surgery (69%) and least among those undergoing plastic surgery (38%).

**Figure 1:**
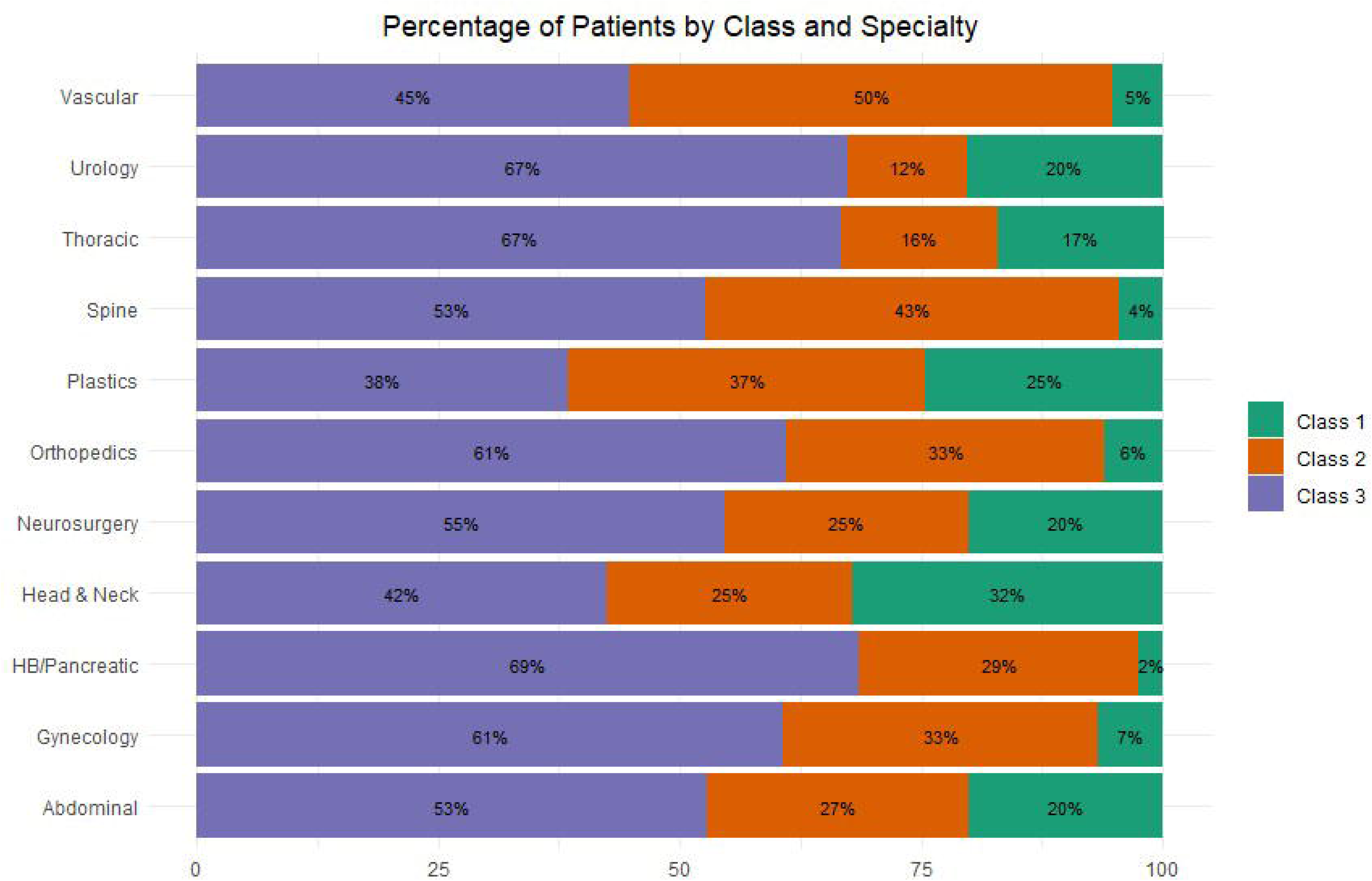
Class Membership by Surgical Category. HB: hepatobiliary

All hospital outcomes of interest were significantly different by class (p<0.001) (Table 3). The proportion of those discharged home was 99% in the LF-HM class, 96% in the LF- LM class, and 77% in the HF-LM class. We observed that the percentage of patients experiencing an extended LOS was highest in the HF-LM class (27%), followed by the LF-LM class (18%), and the LF-HM class (6%). PT and OT visit frequencies were highest in HF-LM class (PT: 2.1 visits/week; OT: 1.1 visits/week), followed by the LF-LM class (PT: 1.2 visits/week; OT: 0.5 visit/week), and the LF-HM class (PT: 0.3 visits/week; OT: 0.1 visits/week).

## Discussion

The results of this study indicate that LCA may be a useful approach for identifying latent subgroups of post-operative older adults, a large patient population vulnerable to poor outcomes. We specifically identified three latent subgroups of post-operative older adults, which we have labeled as Low Frailty-High Mobility (LF-HM) (15.3%), High Frailty-Low Mobility (HF-LM) (27.6%), and Low-Frailty-Low Mobility (LF-LM) (57.1%).

These subgroups were present within each of the included surgical categories, although the proportion of patients belonging to each subgroup varied by type of surgery.

Furthermore, patients in each subgroup differed significantly from one another in discharge disposition, LOS, and utilization of PT and OT services.

The classes identified in this study align with clinical reasoning and highlight the importance of physical function in the International Classification of Functioning, Disability and Health (ICF) domains of mobility and self-care (activities of daily living).^38^ It was not surprising to us that our analysis identified a subgroup of patients (HF-LM) with poorer general health, higher frailty scores, and lower physical function and that these patients were more commonly discharged to post-acute care and experienced higher rates of extended LOS. Likewise, it stands to reason that our analysis identified another class of patients (LF-HM) that were generally healthy, had low frailty scores, had high mobility and daily activity function scores, were nearly always discharged home and rarely experienced an extended LOS. However, less expected was the identification of the LF-LM class, which is similar to the LF-HM group for all class identifiers, with the exception of AM-PAC scores which are both lower in the LF-LM class compared to the LF-HM class. Despite only this difference between groups, the LF-LM class experienced extended LOS rates 3 times higher and utilized 4-5 times as many PT and OT visits per week compared to the LF-HM class. These findings align with previous studies that have highlighted the influence of patient mobility and daily activity function on hospital outcomes and the importance of measuring these items as part of routine care.^17,39–41^

The findings of this study may be useful for the development of tailored postoperative pathways that facilitate more efficient use of hospital services and increase the overall value of postoperative care. For example, we observed that 99% of those in the LF-HM class were discharged home and only 6% experienced extended LOS. On the other hand, those in the HF-LM class were discharged home only 77% of the time and 27% of these patients experienced an extended LOS. In this scenario, it could be argued that services such as PT and OT consults provided to those in the HF-LM class are of greater value, given these patients are at greater risk of undesired outcomes. Similar concepts have been proposed using AM-PAC scores alone, which were shown to have the potential to reduce low-value PT and OT consults among patients admitted to the hospital for medical diagnoses.^42^ This study expands on this approach by examining the influence of multiple patient factors on key outcomes and by identifying patient subgroups that can inform tailored postoperative care pathways.

The classes identified in this study, while corresponding to one institution, are likely generalizable to other institutions performing surgery among older adults seeking to identify clinical subgroups. Importantly, this study was conducted exclusively using routinely collected data extracted from the EHR. While this increases the overall feasibility of the study performed, it does place additional emphasis on the identification and selection of clinical measures, as well as systematic collection and documentation of these measures. Previous publications can provide guidance on the process of selecting outcome measures for healthcare systems seeking to implement or refine data collection strategies.^43,44^

Strengths of this study include a large patient sample that have undergone a wide variety of surgical procedures. This study also utilized data elements that are routinely collected at many hospitals, increasing the generalizability of our results. There are also limitations to this study that should be considered when interpreting results. It is likely that there are factors not measured routinely in our healthcare system that would have altered the subgroups identified in this study had they been included in our analysis. For example, social determinants of health and mental health (i.e., depression) have been shown to influence LOS and discharge disposition among hospitalized patients and may have altered the composition of the classes we identified.^45–49^ Our analysis was also performed specifically among older adults after surgery and it is possible that this affects the generalizability of our results to other populations, such as postoperative adults under 65 years of age.

## Conclusion

We identified three clinically relevant subgroups of postoperative older adults based on data routinely collected in the EHR. We observed significant differences in the proportion of patients discharged to home, the proportion with extended LOS, and utilization of PT and OT services between each of the groups. The design and results of this study may inform the development of more patient-centered care pathways after surgery that maximize patient outcomes and thus the value of postoperative hospital services.

## Author Contributions

KM wrote the primary manuscript draft. AB led the analysis of study data. EH and LG assisted with extraction of clinical data from the medical record. All authors contributed to idea development, provided manuscript revisions, and approved of the final manuscript.

## Data Availability

All data produced in the present study are available upon reasonable request to the authors

